# Using Photovoice to explore HIV self-testing and secondary distribution of HIV self-testing among gay, bisexual and other men who have sex with men in China

**DOI:** 10.1101/2024.06.21.24308294

**Authors:** Qianyun Wang, Ying Lu, Yuxin Ni, Xumeng Yan, Rayner Kay Jin Tan, Dan Wu, Joseph D Tucker, Jason J Ong, Weiming Tang

## Abstract

**Background:** Human immunodeficiency virus self-testing (HIVST) has been globally recognized as a useful and reliable strategy to promote HIV testing, especially among marginalized populations. In order to improve the understanding of HIVST uptake of the test users, notably gay, bisexual and other men who have sex with men (GBMSM) in China, this study aims to qualitatively explore the HIVST uptake experiences among this population.

**Methods:** The study employed Photovoice methodology, theoretically built on documentary photography and critical consciousness, to explore the experiences of HIVST and secondary distribution among GBMSM in China. Participants created photos and citations and shared and discussed them in the group. Three focus groups and one follow-up interview were held virtually for each participant who were self-identified as gay or bisexual and used a self-test kit before.

**Results:** This program recruited 22 participants from all throughout China. The findings revealed that systemic discrimination based on gay identification and AIDS-phobia both inside and outside the gay community, affected participants’ decision to take HIVST and to pass HIVST kits on to others. Participants utilized HIVST on a routine basis, citing sexual health concerns, psychological comfort, and a responsibility to their significant others as reasons for doing so.

HIVST kit *distribution within intimacy,* and *the significant role of gay-led community-based organizations* were found as characteristics of interactions between participants and those who they passed HIVST kit(s) on to. There were observed both facilitators and barriers to HIVST uptake and/or secondary distribution in this demographic.

**Conclusion:** In the study, images and narratives were acquired through empowering GBMSM and promoting their community engagement to underline the necessity for measures and policies on promoting HIVST among this population. Findings also entailed the need to create a more inclusive society for sexual minorities and people living with HIV. Implications for promoting HIVST secondary distribution and limitations and strengths of the pioneer photovoice study among GBMSM in China were also listed.

## Introduction

HIV self-testing (HIVST) has been widely acknowledged as an innovative and reliable method for increasing HIV testing. [1] The World Health Organization (WHO) promotes HIVST as a successful testing strategy, especially for key populations, such gay, bisexual and other men who have sex with men (GBMSM), since it provides better convenience, autonomy, and privacy for test users [2]. In China, despite the high demand for HIVST among GBMSM, previous studies have noted obstacles, including safety, coerced testing, and legality [3], and thus the voice of the end users are essential for planning more sustainable HIVST programs.

The practice of sharing HIVST with others in one’s social network is known as secondary distribution of HIVST (SD-HIVST) [4]. Two studies in China indicated that SD-HIVST among GBMSM can be aided by integrating social media, monetary incentives and peer referral [5,6].

A few qualitative studies have investigated HIVST experiences among GBMSM in China. For example, it was discovered that HIVST empowered GBMSM and informed sexual decision-making, based on qualitative data collected from interviews with 42 GBMSM and 6 stakeholders. [7]. A recent study conducted in-depth interviews with 22 GBMSM who had participated in secondary distribution of HIVST in southern China, and found the facilitators and barriers to HIVST use for this population [8].

While more studies have begun to focus on HIVST and SD-HIVST among GBMSM in China, there has not been research using qualitative methods to dig deeper into issues like how social interaction and/or cultural norms may affect GBMSM’s decisions regarding HIVST and SD-HIVST or employing a participatory action research (PAR) tool to engaging GBMSM for the outcomes with their both collective and self-reflective voices.

Photovoice, a community-based PAR approach, has been viewed as a technique for empowering disadvantaged people in which participants utilize a combination of photography and critical discussion to highlight their community’s issues and demands for social change [9]. Photovoice has been used in a rising number of studies to enable community dialogue and advocacy in the fields of health and public health, with a focus on people living with HIV (PLWH) [10–12].

This study aims to fill the gap by using Photovoice method to qualitatively explore the experiences on HIVST and secondary distribution of HIVST from the perspective of GBMSM in China. Through the process, we intent to inform understandings, as well as collective and self-reflective voices from the GBMSM community. This study will answer the questions:

1. What are the GBMSM’s own awareness of and/or attitude towards HIV/AIDS and HIVST?
2. What are the characteristics of interacting relations between index participants and alters during HIVST secondary distribution?

## The Theoretical Framework

Photovoice methodology is theoretically built on documentary photography, feminist theory, and critical consciousness. Documentary photography aims to use visual narratives to capture the social lives of a specific group of people in time and space, and Documentary photographers always concern about social equity and use comprehensive content to help raise social awareness and make a difference [13,14].

The principles of feminist theory lie in that the personal is political and no one is better to study and understand the issues of a group than the people within the group [15]. Although this theory was developed as a response to women’s social experience, its principles are also applicable to others [12]. In many cases in China, the lack of GBMSM’s uncovered experiences lies in their marginalized political identity as gay men. In this study, participants were believed to identify their community issues and needs, such as their experiences of conducting and distributing HIVST, from their perspective.

Critical Consciousness is grounded where community involvement can lead to social change and social equity, and it encourages community residents to have critical group dialogue in order to foster critical understanding and action for community change [16,17].

## Methodology

### Ethics

We registered this trial on the Chinese Clinical Trial Registry website (ChiCTR2000039632). This study received Institutional Review Board approval from the Dermatology Hospital of Southern Medical University (2019020[R3]). Oral consent was obtained from all participants. In anticipation of the inaugural virtual focus group session, each participant was furnished with an electronic consent form well in advance. This facilitated ample opportunity for them to comprehensively peruse the associated content. Should they express their willingness to partake in the proceedings, a designated ZOOM meeting link was provided, enabling their presence at the inaugural focus group session. Commencing the inception of this session, the researcher reiterated the contents of the consent form, subsequently inquiring into the participants’ continued willingness to engage in the program, with participation proceeding for those who chose to remain within the group.

### Recruitment

The study conducted from June to August, 2021, was part of a quasi-experimental trial which explored the effectiveness of sexual health influencers identified by an ensemble machine learning model in promoting SD-HIVST among GBMSM in China [18]. The Social Entrepreneurship to Spur Health team, the Zhuhai Center for Disease Control and Prevention, and the Zhuhai Xutong Voluntary Services Center, a GBMSM community-based organization (CBO), collaborated in this study.

The recruitment criteria for this photovoice are: 1) 18 years and above; 2) was assigned male at birth; 3) ever had sex with men; 4) ever conducted HIVST; 5) ever distributed HIVST or was planned to do so; 6) have a device (e.g., mobile phone) to take photos and know how to use it. In this study, GBMSM from different cities in China virtually gather to share experiences and thoughts as a process of community participation. Individuals were excluded if they were unable to attend three focus group discussions or take a follow-up interview. An incentive of RMB 150 (USD 20) was provided to each participant in three equal portions in the first, second, and third focus group.

### Sampling

Purposive sampling was used to recruit participants. One participant was recruited by snowballing. He was informed with this project by another participant, his friend. He approached the research team and expressed his willingness to participate in the project. After examining that he met all the recruitment criteria for participation, the research team confirmed his participation.

### Data Generation

Each focus group discussion section was co-facilitated, once a week, by three research assistants from SESH team. The study procedures are presented in Figure 1.

**Figure 1.**
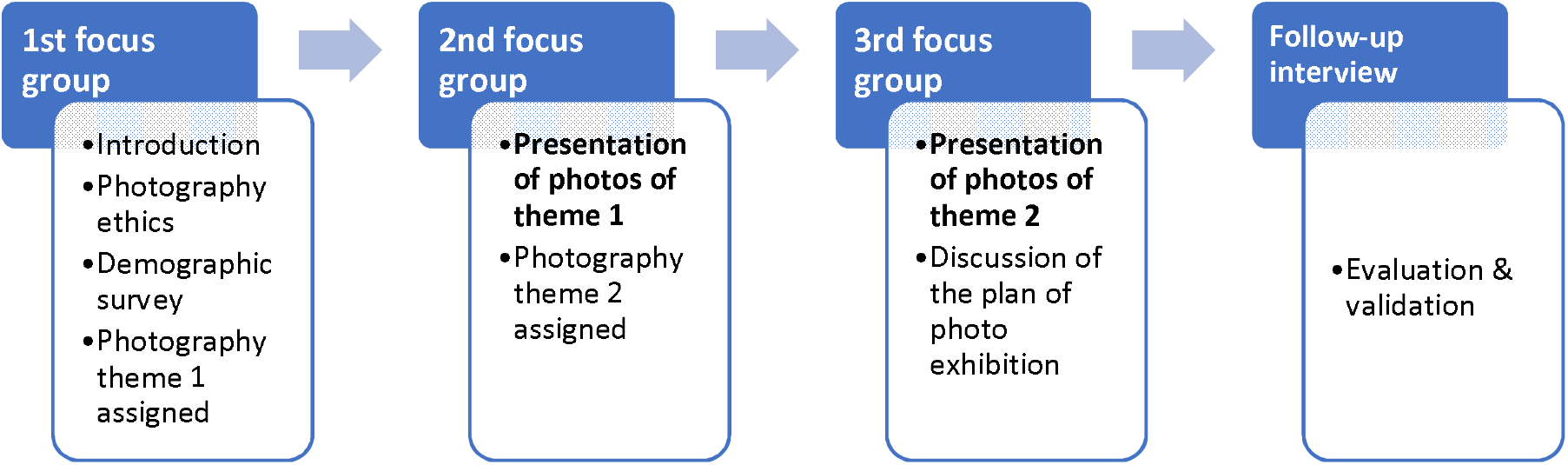
Study Procedures

The first focus group aimed at discussing the goal of the study, the introduction of Photovoice procedure, and ethics of photography. In the first focus group, participants filled in a demographic survey. This questionnaire only collected overall profiles, each data could not be matched to an individual) and were asked to take three to five photos to answer the following questions: *“What are the phenomena related to HIV/AIDS in life?”; “How do you feel about HIV self-testing? What is your take on it, your attitude toward it, or your experience with it?”*.

Participants shared and explained their images and captions during the second focus group, which took place one week later. After the group discussion in the second focus group, participants were then instructed to take three to five more images in order to respond to the following questions: "What are your ideas, feelings, and experiences with secondary HIV self-testing?"; "Please explain the person or scenario to whom you provide the HIV self-test kit(s)," which were presented and interpreted in the third session after another week.

### Data Analysis

Each group discussion was audio-recorded and transcribed. Photos were collected as visual data. The research team reached preliminary results with the strategy proposed by Tsang [19]. The results were shared with participants individually during the follow-up interviews, which were also recorded. A four-step Photovoice data analysis strategy proposed by Tsang [19] was used, as follows:

1. Photograph Analysis Based on the Researcher’s Interpretations: In step 1, a preliminary explanation of the phenomenon was created by the study team after conducting an initial analysis of the photographs. We divided them into categories and looked at each one in detail, then divided them again and looked at the categories again until saturation was reached.
2. Photograph Analysis Based on the Participants’ Interpretations: Following a preliminary examination of the photographs and the development of the researcher’s explanation, we discovered whether there were any other possible explanations for the photographs based on the participants’ interpretations. Therefore, we examined the photographs’ descriptions and as well as the data from the focus group discussions.
3. Cross-comparison: Then, in order to develop an integrative explanation of the phenomenon based on the researcher’s interpretation and that of the participants, we compared photographs in terms of content, narratives, themes and others.
4. Theorization: Last, we generated narrative and visual representations and explanations of the phenomenon based on the analysis from the cross-comparison.

Participants’ meanings were verified using narrative data from follow-up interviews by one researcher. Each of these factors are described with illustrations drawn from the transcripts of focus group discussion and photos with captions, denoted by participants’ pseudonym. All names mentioned in this paper are pseudonyms.

## Results

This study recruited 22 participants from 5 provinces in China. The ages of the participants ranged from 19 to 50, with an average of 28 years old. Eighteen individuals (82%) self-identified participants self-identified as gay, three (14%) said they were bisexual, and one (4%) reported not sure or did not want to share. These specific identifying details were gathered without being linked with any particular participant.

Findings highlights the stigma and discrimination connected to HIV and HIVST within and outside of the community of GBMSM, HIVST as a daily-routine behavior among participants, interactional characteristics during HIVST secondary distribution.

### Stigma and Discrimination Connected to HIV

During the focus groups, stigma and discrimination towards HIV was heavily discussed, and some participants offered photos and captions reflecting their views toward and/or understanding of HIV.

### Social Stigma in HIV Attached to the Gay Identity

For example, Stephen described how he felt about the HIV stigma associated with his identity, stating, as a sexual minority:

“We [as gays] often suffer from two diseases [stigma] at the same time - sexual metamorphosis and HIV. Homosexuality and HIV [status] are ‘covered up’ in reality. Society wants us, and we sometimes want, to hide.”

**Image and Caption 1.** (The photo has been removed due to the inclusion of Chinese language content. Please contact the corresponding author to request access to this information.) A cover of the book *Covering: The Hidden Assault on Our Civil Rights* **by Stephen**

In the set of two photos of Stephen’s roommate’s blood donation certificate, Stephen shared his concerns that he would never be accepted to donate blood as gay. Officially, male gays are not permitted to donate blood in China. However, because Chinese sexual minority identities are frequently hidden, this formal law has not been implemented. As a result, it represents China’s systemic stigma towards the community of GBMSM.

**Image and Caption 2.** (The photos have been removed due to the inclusion of Chinese language content. Please contact the corresponding author to request access to this information.) “Roommate’s blood donation certificate” **by Stephen**

### “AIDS” Phobia within the Gay Community

Many of the participants’ stories revealed their fear of HIV. Participants in several situations voiced their irrational fear of contracting HIV. Xiao Li, for example, when discussing gays’ “AIDS” phobia, which is termed as “Kong Ai” in Chinese, said:

“Some people [gays] will be so afraid, that is, they feel they are infected with HIV, but they are not. Then they will always feel they are, always be afraid, and then [mentally] worse than those who are actually living with HIV.”

Particularly, Pando pointed out that his “AIDS” phobia might have contributed to his use of HIVST, as he quoted:

“To be honest, I think I should also be one of those who fear AIDS. I often do HIV [self] testing. I pay great attention to this kind of things [HIVST].”

### Personal Transformative Development on Anti-Discrimination in HIV

Although mixed with the fear of getting infected with HIV and perceived discrimination in HIV targeting gays, updated concepts related to HIV were included in focus groups; for instance, “U=U” (Undetectable equals Untransmutable) was advocated by some participants.

Some also presented their personal transformative development on anti-discrimination in HIV. They shared that the friendship with other gay friends who were living with HIV helped them dispel the myths about HIV:

**Image and Caption 3.**
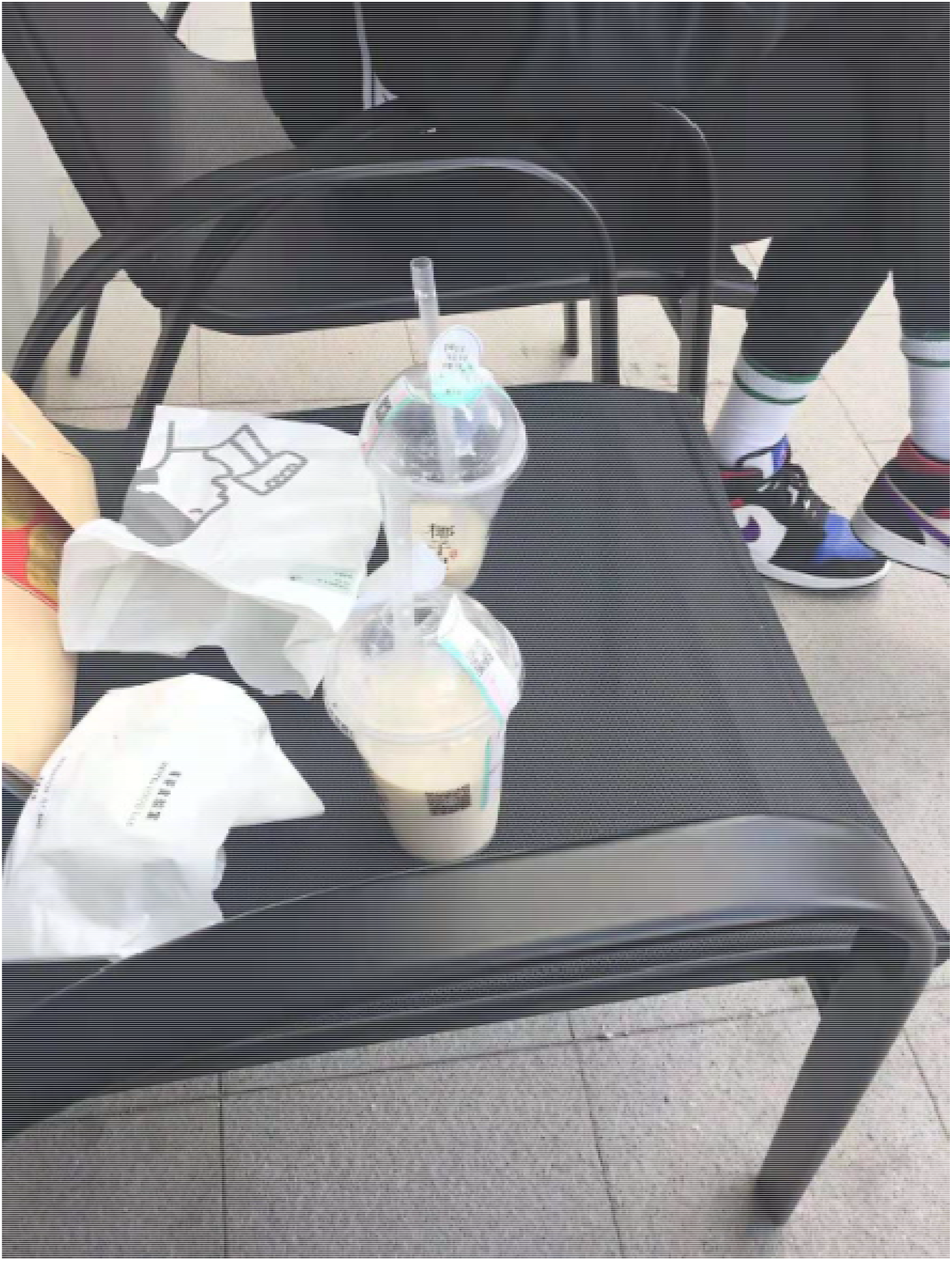
“The friend beside me is diagnosed with HIV, but we often hang out for food. It turns out that it is absolutely not contagious through daily contact.” **by Carey.**

Additionally, Lao Wang shared that HIV-related education and knowledge transferred his understanding of HIV which was shaped in his childhood:

“The generation like my mother’s say it [AIDS] is a terrible disease. Don’t have meals or live with people living with HIV. My mother spoon-fed me with that concept. Well, then, when I went to college, I began to know some knowledge given by our school and some lectures about sexual health. In fact, it was not that terrible.”

Specifically, one participant, Ririyeye, summarized three stages of understanding HIV among gays, from a developmental perspective:

“1^st^ : People are ignorant of HIV. Though there are only a variety of news and reports on HIV cases, HIV is perceived to be far away from daily life.

2^nd^ : Fear emerges when it comes to HIV. After an ignorant stage, people believe HIV is the most terrible disease, where daily contact will lead to infection. HIV is impossible to prevent, and with no medicine to cure it, even people living with HIV die young.

3^rd^ : HIV has been simplified [normalized]. HIVST kits are accessible. There is PrEP for early prevention, PEP for blocking, and medical adherence can also lead to a normal life.”

## HIV Self-Testing as Routine

Participants shared that they utilized HIVST on a routine basis, citing sexual health concerns, psychological comfort, and a responsibility to their significant others as reasons for doing so.

### Sexual-Health-Oriented Decision

Almost all participants agreed that their decision to use HIVST was based on sexual health. They identified the significance of knowing one’s own HIV-status, for a better linkage to care if necessary. In that way, HIVST served them well due to the greater convenience, autonomy and privacy it offered.

**Image and Caption 4.** (The photo has been removed due to the inclusion of Chinese language content. Please contact the corresponding author to request access to this information.) “HIV Self-testing in the dormitory on campus at night. --Regular self-testing released my fear [of infection]. Do not fear, do protect your body, do early tests and early treatment”. **By Xiaoli**

Further, Qingfeng said HIVST became a part of his daily life:

“Now I’m applying for HIVST from [name of a community-based organization (CBO)], and it is free. I apply for it every month. Self-testing has become a part of life.”

### Psychological Comfort

It was also expressed that HIVST sometimes was served for psychological comfort by several participants. As Caihong explained,

“Personally, I may have sex no more than twice a year. So, it [HIVST] is more out of curiosity than the test results.”

Furthermore, Ririyeye stated that regular self-testing might help alleviate his “AIDS” phobia by confirming his negative status on a regular basis. As his work presented,

**Image and Caption 5.**
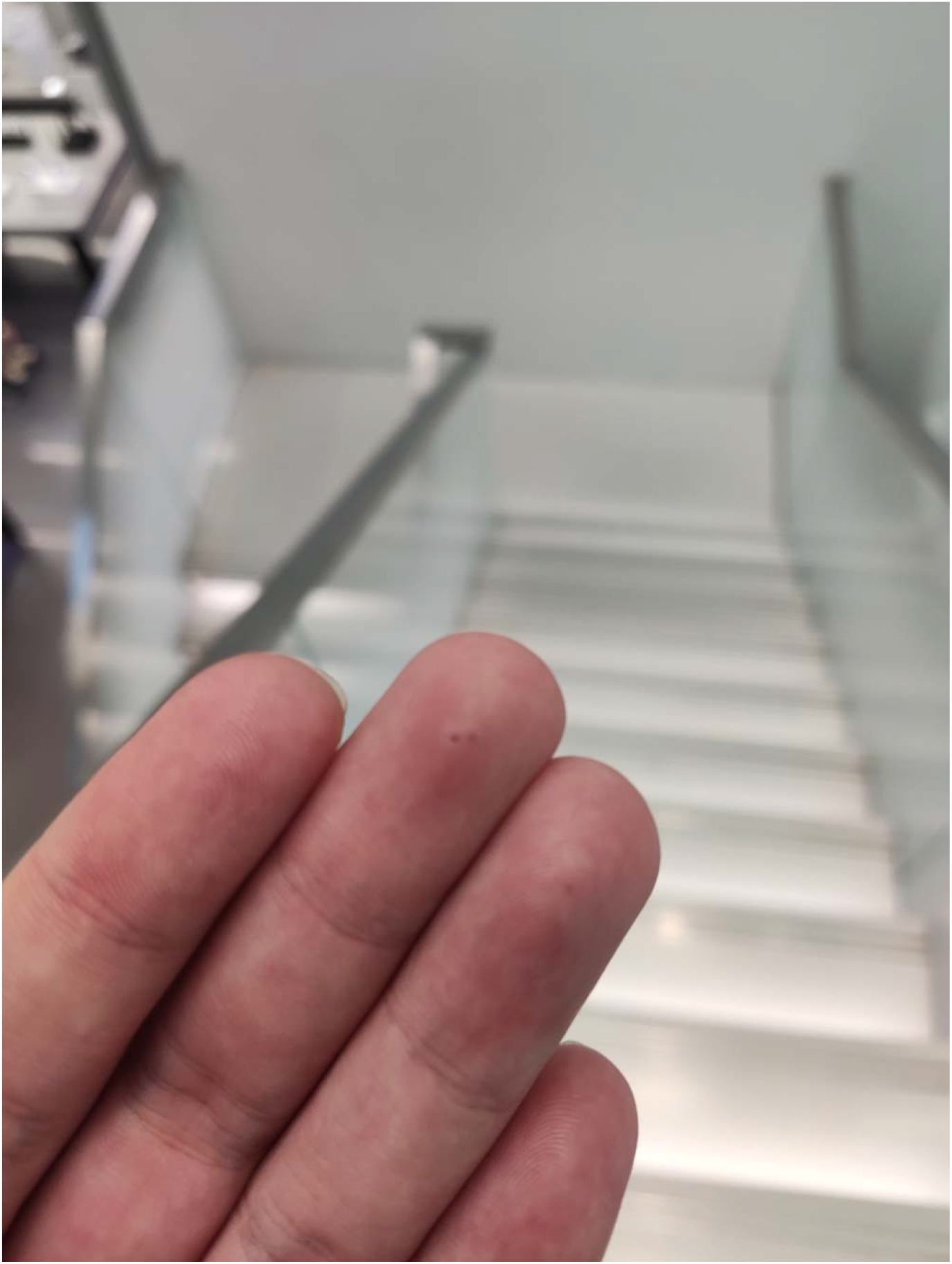
“Frequent self-testing brought me three pinholes. That is my anxiety”. **By Ririyeye**

### Moral Duty for Oneself and Others

It was discovered that taking HIVST on a regular basis was also motivated by moral obligation, both for oneself and for others, such as family and friends.

"It is responsible for family and friends to take regular self-testing," Carey Zhang, for example, said. And he further explained,

“If you take medicine in time [after being tested positive] well, you can ensure your life span and your quality of life are of no different from those of normal people. Therefore, in that way, I think, to ensure my quality of life and living condition is to be responsible for my family and friends.”

## Interactional Characteristics during SD-HIVST

### A Collective Sense of the Gay Community

When some participants described their understanding of HIVST and SD-HIVST in the gay community, they provided a collective sense where they felt all the residents were connected with each other in terms of sexual health, like the underground tree roots, or stars in a galaxy.

Promoting HIVST uptake in the community is beneficial to everyone.

**Image and Caption 6.**
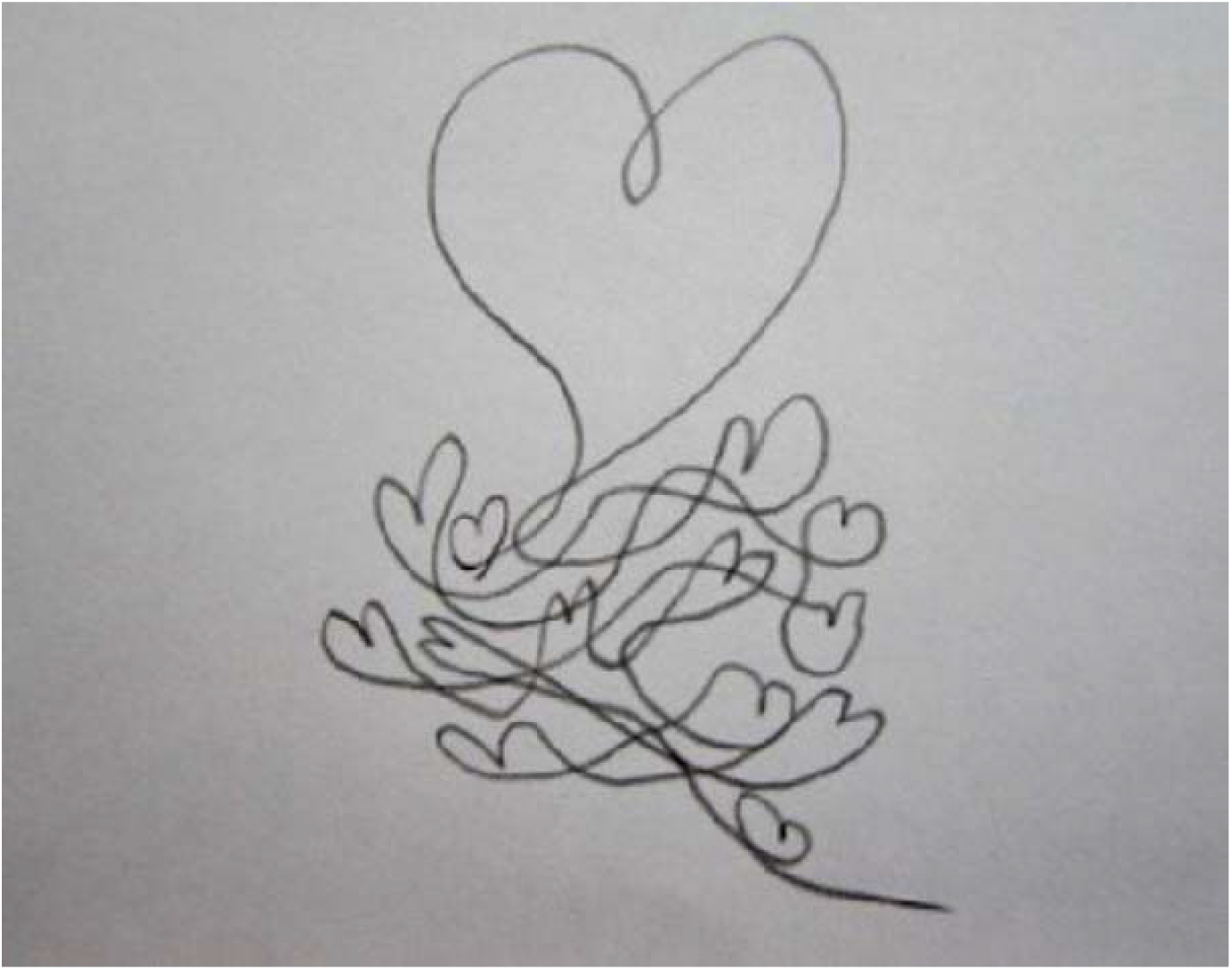
“My own painting – our [we and alters] hearts are connected.” **By Qian**

Further, Stephen pointed out how he expected HIVST promotion in the community to help eliminate the systematic discrimination against the gay population:

“It is hoped that through promoting HIVST uptake, the HIV pandemic can be better controlled, and discrimination and stigmatization [related to HIV against gays] can be eliminated gradually”.

Similarly, participants distributed HIVST kits, in many cases, to those who were also from the gay community. And they viewed their reciprocal relations as a prominent factor for the self-driven distribution:

“HIVST secondary distribution is voluntary, reproducible and transmissible. The stars can light up the sky, when all come together; it turns out to be a galaxy of group immunity.” **By Qian**

### ‘More Than just Online Friends’: Necessary Face-to-Face Interaction

Almost all the participants agreed that the gay dating app was an important approach for them to develop their networks for friendship and/or intimacy among this population. Namely, their social networks with other gays have an internet-based connected face. Furthermore, convenient shipping systems in China enable online secondary distribution. However, when it came to the persons whose participants distributed HIVST to, participants usually prioritized those who had face-to-face interactions with them before, such as gay friends whom they met in a CBO, schoolmates, and sexual partners, among others.

For example, Stephen shared a smart phone screenshot where he had a chat with an online gay friend in which he was attempting to send out an invitation of online application for HIVST but failed. However, he then offered the invitation to a schoolmate. HIVST distribution, he argued, should be based on trust, which he said could not be guaranteed by just online pals.

**Image and Caption 7.** (The photo has been removed due to the inclusion of Chinese language content. Please contact the corresponding author to request access to this information.) [translation -“Hey there.” -“?” -“Nothing.”] “In the beginning, I thought there were many people who could be the recipients of the distribution, like online friends on WeChat, strangers on the dating APP, or people you’ve only met briefly. But as I started to distribute, I realized that I ended up choosing the most understanding friend.” **By Stephen.**

Many other participants were able to relate to Stephen’s experiences with HIVST distribution. As Haiqing explained as followed:

“The reason why I didn’t pass it [HIVST kits] [to the online friends] is that I didn’t know where their boundary was. If I was not familiar with them enough, I would be so embarrassed [to start a HIVST distribution conversation].”

This occurrence was linked by some participants to HIV stigma and/or AIDS fear, which was prevalent in the gay community.

“I think to conduct a distribution is difficult [among gays]. Some gays will be just afraid of HIV testing, especially those who had high-risk sexual behaviors.” **By Lao Wang** “[If I want to pass the kit to an internet person], he will wonder why I want to pass this thing to him……It’s possible that he thinks you have bad intentions. Your motivation to trigger this invitation is embarrassing. He might assume that you think he is ill [HIV positive].” **By Pando**

### HIVST Secondary Distribution with Intimacy

Specifically, some participants shared they passed HIVST kits to sexual partners. It was common that distribution happened right before their sex.

“I would for sure ask him [the sexual partner] to self-test [right there]. Of course, safe sex must be taken. If we still have a connection after sex, I would certainly like to pass [online] distribution to him. From my point of view, I can also take a test, which can also indirectly prove that our previous sex was safe.” **by Pando**

Lichee presented his experiences of HIVST and secondary distribution to his boyfriend when they were just committed to their romantic relationship. He thought it was fair to know each other’s health status at the beginning.

**Image and Caption 8.** (The photo has been removed due to the inclusion of Chinese language content. Please contact the corresponding author to request access to this information.) “Buy one [HIVST kit] get one for free.” **By Lichee**

“Then I told him about the test, but he questioned whether I thought he was hooking up or cheating. But in fact, after I explained to him later, he also agreed [to have HIVST with me].” **By Lichee**

### The Significant Role of GBMSM-Led CBO

GBMSM-led CBO were described as a platform for interaction in the gay community.

Not only did participants receive services and information from GBMSM-led CBOs, where they presented that they got HIV prevention knowledges, counseling services, free HIVST, etc. For instance, Wind commented that he accessed free HIVST kits from a CBO, and it had become a regular event:

“I now apply for HIVST kits [from a CBO]. They are free. I apply every month [and do self-test] as a part of my life, a fixed part. Now I keep my mind to do self-tests.” **By Wind.**

“It let me know that volunteers [from GBMSM-Led CBO] are empathetic and passionate about promoting of HIV prevention knowledges, which has effectively increased my understanding.” **By Xiao Li.**

Also, some participants were working as volunteers and full-time employers in CBOs, and they had been involved in HIVST and/or other sexual health-related promotion work.

**Image and Caption 9.** (The photo has been removed due to the inclusion of Chinese language content. Please contact the corresponding author to request access to this information.) “On [a date], volunteered to hand away free lubricants and condoms near a gays-gathering-place.” **By Rainbow**

Xiao Zhi, a full-time staff working in a GBMSM-Led CBO, shared his experience:

“We regularly organize HIV-related knowledge training among high school students, college students and other youths, including HIVST kit training. So that we can better understand this group [male gay community], understand HIV, [the way to] protect ourselves……Our goal is to improve the awareness of self-protection, know and understand AIDS, not fear AIDS, and strengthen sexual minorities.”

GBMSM-led CBOs efficiently promote HIVST uptake among GBMSM and become an engaging platform where they could have community participation through volunteering and working with payment.

## Discussion and Implication

Our discussion and implication include 1) stigma and discrimination working as an essential barrier of HIVST as well as secondary distribution, 2) the individual factors contributing to usage of HIVST, while the collective factors contributing to secondary distribution, and 3) strengths and limitations of this Photovoice project on GBMSM in China.

### Stigma and Discrimination affecting HIVST and Secondary Distribution

We found that stigma and discrimination related to HIV/AIDS and gay identity would influence Chinese GBMSM’s HIVST behaviors directly and indirectly. GBMSM discussed how they felt stigmatized as a sexual minority in terms of HIV/AIDS by other individuals. By unleashing the fear of HIV/AIDS infection, focus groups frequently expressed “AIDS” phobia. Certain participants spoke of the mental load associated with the possibility of acquiring HIV/AIDS. Excessive HIVST uptake may be a behavioral reflection of this mental anxiety, which might encourage excessive and unnecessary HIVST behaviors among this population.

Stigma and discrimination connected to HIV have been found linked to an increase in risk-taking behavior and a decline in effective HIV prevention and testing among these males [20]. According to US studies, GBMSM who internalized stigma as gay men as were less likely to conduct HIV testing and more likely to take sexual risk behaviors [21,22]. However, previous research, which examined the relationship between stigma and discrimination and HIV testing behaviors, did not specifically address HIVST. Further research should concentrate on exploring elements impacting this population’s mental health, particularly, stigma and discrimination linked to HIV/AIDS and gay identity, and how these could further effect HIVST behaviors.

Stigma and discrimination connected to HIV have been found linked to an increase in risk-taking behavior and a decline in effective HIV prevention and testing among these males [20].

According to US studies, GBMSM who internalized stigma as gay men were less likely to conduct HIV testing and more likely to take sexual risk behaviors [21,22]. However, previous research, which examined the relationship between stigma and discrimination and HIV testing behaviors, did not specifically address HIVST. Further research should concentrate on exploring elements impacting this population’s mental health, particularly, stigma and discrimination linked to HIV/AIDS and gay identity, and how these could further affect HIVST behaviors.

The study also found that stigma linked to HIV/AIDS and “AIDS” phobia among GBMSM could affect HIVST secondary distribution, and the persons to who they distributed HIVST kits. In the study, it was common that HIVST secondary distribution happened between sexual partners right before their sex. However, due to participants’ stigma around HIV/AIDS, it might also be unsettling to ask sexual partners to conduct HIVST. Despite the heavy use of online dating apps and other social networks, and the accessible HIVST online delivery services, participants still disagreed that online networks could facilitate SD-HIVST. Considering trust issues and a fear of being stigmatized associated with HIV/AIDS, even if they had extra kits, some GBMSM said that it was challenging for them to start an online conversation about HIVST kit distribution; instead, they placed more importance on giving out kits to reliable individuals, including friends with whom they engaged offline. Secondary distribution of HIVST might be hampered by “AIDS” phobia and stigma among GBMSM, both through online and on-site social networks. From a social network perspective, further research should explore how stigma and discrimination around HIV/AIDS among GBMSM can influence their behavior of HIVST and secondary distribution.

In the study, participants’ transformational anti “AIDS” phobia development experiences were uncovered. They shared that they benefited from their social network in the process of eradicating the stigma associated with HIV and AIDS and in learning new information about HIV prevention. Furthermore, HIV stigma reduction efforts should engage various public sectors in addition to relying on one’s personal network. A study from Ghana modified HIV stigma reduction training for GBMSM using a mixed methods approach. They discovered that addressing HIV and other intersectional stigmas related to sexual and gender diversity was the fundamental condition for eradicating stigma [23]. The outcomes are consistent with our findings that GBMSM experienced many forms of stigma, such as those related to HIV and homosexual identity. The application of HIV stigma elimination in practice should be improved by future research.

### Individual and Collective Factors for HIVST and Secondary Distribution

Most participants mentioned that because of the convenience and privacy it offered, self-testing has become a regular health check habit. Other than China, a few studies also found similar feedback from GBMSM that HIVST novel characters other than facility-based testing contributed to more HIV testing. According to focus group discussions (FGDs) conducted in Uganda with 74 GBMSM and 18 health workers, GBMSM preferred HIVST to facility-based HIV testing procedures because of its convenience and confidentiality [24]. Another study from the UK used focus group discussions among 47 GBMSM and found that the convenience of use and confidentiality of HIVST led to testing. However, there were also issues raised about the window period, overly complicated HIVST kits, and other factors [25].

In comparison to the facility-based method, the HIVST safeguards individual sense to a larger extent, namely “the right to privacy” [26]. Most participants who discussed their own usage of HIVST said that because of the convenience and privacy it offered, self-testing has become a regular health check habit. In comparison to the facility-based method, the HIVST safeguards individual sense to a larger extent, namely “the right to privacy” [26].

While HIVST has been credited with defending “the right to privacy” for self-testing individuals, it has also highlighted secondary distribution as one of the most efficient means of promoting HIVST [27]. At this point, the collective sense may thus be crucial. For instance, several participants in the study identified as belonging to the community [of gay men], and they thought that when HIVST and secondary distribution was collectively practiced in the community, it would promote community health and help community members overcome the stigma associated with AIDS.

The roles of both individual and collective cultural symbols in GBMSM HIV testing were examined in a study conducted in the Philippines [28]. Filipino GBMSM collective culture shared inside and outside the GBMSM community, including family, religious communities, helped the population cope with HIV, depression connected to HIV, stigma, and other issues. Yet, the study in the Philippines was unable to explore how individual and collective cultures would affect HIVST and secondary distribution. Our study added to the gap by finding that while HIVST was mostly driven by the individual sense, secondary distribution of HIVST was usually motivated by the collective sense. We also observed an association between HIVST and the collective sense, as some participants claimed that knowing their own HIV status—as confirmed by HIVST—also made them feel responsible for their friends and family. Even yet, the study in the Philippines was unable to explore how individual and collective cultures would affect specifically HIVST and secondary distribution.

Furthermore, although HIVST maximized privacy for GBMSM to have HIV testing, the study indicated that community participation encouraged GBMSM to use HIVST as well as secondary distribution. GBMSM-led CBO played an essential role, whereby taking part in collective events, GBMSM were informed about HIVST and given access to HIVST kits. In return, some GBMSM also volunteered to promote HIVST and HIV prevention knowledge in the organizations. Secondary distribution happened in a community level. Community-based approaches have been recognized as effective involvement of HIVST promotion, as various stokeholds together reached community engagement [29,30]. Public sectors and academia should work closely with community-based organizations, when involving with GBMSM for HIV prevention. Furthermore, although HIVST maximized privacy for GBMSM to have HIV testing, the study indicated that community participation encouraged GBMSM to use HIVST. GBMSM-led community-based organizations played an essential role, whereby taking part in collective events, GBMSM were informed about HIVST and given access to HIVST kits. In return, some GBMSM also volunteered to promote HIVST and HIV prevention knowledge in the organizations. Community-based approaches have been recognized as effective involvement in HIVST promotion, as various stokeholds together reached community engagement [29,30].

Public sectors and academia should work closely with community-based organizations, when involved with GBMSM for HIV prevention.

To more properly promote HIVST usage and secondary distribution, we need to pay attention to the function of individual and collective senses, which could be balanced and complementary. Researchers or other public health sectors may carry out more research in the area.

### Strengths and Limitations

This study is an innovative use of Photovoice in the GBMSM community in China. First off, the community does not widely employ Photovoice approach. During recruiting, the researchers spent a significant amount of time explaining photovoice and fielding numerous inquiries from community members who were interested. Recruitment and participation might be difficult due to participants’ unfamiliarity with the activity approach, but this is a fantastic example of experimenting. Nearly all respondents to the follow-up survey said that they would want to see and participate more community-based PAR programs.

Secondly, it’s important to note that this Photovoice project was exclusively performed online because to the COVID epidemic restriction, making it more inclusive in time and space and attracting a diversity of participants in terms of age and geographic location. However, there were stricter guidelines for participants’ usage and accessibility of technological devices.

Additionally, digital elements endured throughout the project, e.g., some participants would directly reply to an interactive scenario in the submitted photos by taking screenshots of their mobile phone conversations, etc. An online photographic exhibition was also planned in order to increase the impact of the research findings. The limitations and benefits of digital PAR should also be further studied in future study.

Many empowering events occurred throughout the participants’ exploration, such as the participants’ own definition of the inequalities and the systemic exclusion they personally experienced related to their sexual minorities identity, asking for the eradication of such from all spheres. Some participants also shared more updated HIV prevention messages, e.g., U=U, to dispel the overwhelming fear of HIV and AIDS among others in the group. Nearly all participants in the event reported, in the follow-up survey, gaining new meaningful knowledge. Photovoice enables participants themselves addressed the issues they confront as community members. As a discursively excluded minority in Chinese society, GBMSM ought to have more chances to speak up for their communities and themselves.

The cycle period for this Photovoice project was relatively short. Within a week, each participant was asked to contribute photographic images and citations in response to several themes, which were then shared with the group. Some claimed that they were so hurried that they did not share their work on time. An extended project cycle may ensure participants’ perception of involvement and experience.

## Data Availability Statement

The data that support the findings of this study are available on request from the corresponding author, Weiming Tang. The data are not publicly available due to ethical restrictions and privacy concerns, as they contain potentially sensitive information that could compromise the privacy of research participants.

